# Low rates of enteric pathogenic bacteria and resistance gene carriage in the sheltered homeless population in Marseille, France

**DOI:** 10.1101/2020.09.22.20199729

**Authors:** Tran Duc Anh Ly, Linda Hadjadj, Van Thuan Hoang, Ndiaw Goumbala, Thi Loi Dao, Sekene Badiaga, Herve Tissot-Dupont, Philippe Brouqui, Didier Raoult, Jean-Marc Rolain, Philippe Gautret

## Abstract

We aimed to assess the prevalence of pathogenic bacteria and resistance genes in rectal samples collected among homeless persons in Marseille, France. In February 2014 we enrolled 114 sheltered homeless adults who completed questionnaires and had rectal samples collected. Eight types of enteric bacteria and 15 antibiotic resistance genes (ARGs) were sought by real-time polymerase chain reaction (qPCR) performed directly on rectal samples. ARG-positive samples were further tested by conventional PCR and sequencing. We evidenced a 17.5% prevalence of gastrointestinal symptoms, a 9.6% DNA-prevalence of enteric bacteria carriage, including *Escherichia coli* pathotypes (8.7%) and *Tropheryma whipplei* (0.9%). Only 2 persons carried *bla*_CTX-M-15_ resistance genes (1.8%), while other genes, including carbapenemase-encoding genes and colistin-resistance genes, (*mcr*-1 to *mcr*-*6, mcr-8*) were not detected. Our results suggest that sheltered homeless persons in Marseille do not have a high risk of harbouring gastrointestinal antibiotic resistant bacteria.

## Introduction

Little information is available about gastrointestinal bacterial infections in homeless populations. During 2015–2016 a multistate outbreak of *Shigella* occurred among homeless persons in Oregon, USA. There, the homeless accounted for half of cases [1, 2]. *Shigella* was also responsible for an outbreak occurring among homeless persons and healthcare workers in a homeless shelter in British Columbia, Canada, in 2015 [3]. A survey conducted in Georgia, USA in 2018 reported a high prevalence (23%) of enteric pathogens in homeless individuals’ stools open-defecated on the street, including enterotoxigenic *Escherichia coli* (12%) and *Salmonella spp*. (3.8%), posing health risks to the general public [4]. Other than these enteric bacteria, *Mycobacterium tuberculosis* (causing gastrointestinal tuberculosis), hepatitis A virus and many intestinal parasitic infections have been described in homeless populations [5-10].

Given the lack of surveillance due to the high mobility of this population, antimicrobial resistance (AMR), if occurring in the homeless population, can challenge local health care systems. Production of specific inactivating enzymes (such as extended-spectrum β-lactamases [ESBLs] and carbapenem-hydrolysing β-lactamases) is considered the most important mechanism contributing to antimicrobial resistance to β-lactam antibiotics in gram-negative bacteria [11]. Colistin is currently prescribed as one of the last-line antibiotics for treatment of a variety of human infections; nevertheless, the emergence of plasmid-mediated colistin resistance genes, such as the *mcr*-1 gene, has also been globally observed [12]. Few studies are available regarding the prevalence of enteric pathogens resistant to antibiotics in this population. A high prevalence (75%) of *bla*_CTX-M-15_ was evidenced in 36 ESBL-producing *Enterobacteriaceae* isolated from stools of Tanzanian street children that were phenotypically resistant to tetracycline (100%), trimethoprim-sulfamethoxazole (97%), ciprofloxacin (69%) and gentamicin (44%) [13].

Surveys have been conducted by our institute among homeless persons within two shelters (A and B) in Marseille, France, between 2010 and 2011 and showed a high prevalence (12.9%) of *Trpheryma whipplei* in homeless persons’ stool samples [14]. Gastrointestinal infections were diagnosed in 6% of hospitalised homeless persons at the infectious disease units in Marseille, France between 2017-2018[15]. In a previous work, using direct molecular detection, we observed a lower prevalence of resistance genes in nasal swabs in sheltered homeless in Marseille when compared to a non-homeless population [16]. In this cross-sectional study, using the same approach, we aimed to assess the prevalence of several gastrointestinal pathogens DNA and resistance genes carriage in the sheltered homeless in Marseille.

## Materials and Methods

### Ethics approval and informed consent

This protocol was reviewed and approved by the Marseille Institutional Review Board/Ethics Committee (Homeless population: 2010-A01406-33; Comparison group: 07-008-IFR 48). Informed consent was dated and signed by all individuals.

### Study design and sample collection

Data and rectal swab samples were obtained from adult homeless persons living in two municipal emergency shelters A and B in Marseille, France on 11-13 March and 11 April, 2014, respectively. The participants were asked to answer a questionnaire, including information on demographics, personal history, clinical gastrointestinal symptoms, including diarrhoea (defined as at least three loose or liquid stools per 24 hours), vomiting, nausea, constipation and abdominal pain. Rectal swab samples were collected and stored as previously described [17].

### DNA extraction

Semi-automated DNA extraction was performed on 200 µl of each sample as previously described [18] using a BioRobot®EZ1 Advanced XL instrument (QIAGEN, Hilden, Germany) and DNeasy® Blood & Tissue according to the manufacturer’s instructions. The DNA extraction quality was assessed by RT-PCR targeting internal control TISS phage that was added to each extraction [19].

### Real-time PCR

#### Identification of enteric bacteria

A multiplex PCR-based assay using LightCycler®480 Probes Master kit (Roche diagnostics, France, according to the manufacturer’s recommendations) was used to determine the presence of the *ipaH* gene of *Shigella spp*./EIEC (enteroinvasive *E. coli*), stx1 and stx2 genes of enterohaemorrhagic *E. coli* (EHEC), EAF and EAE genes of enteropathogenic *E. coli* (EPEC), pCVD432 gene of enteroaggregative *E. coli* (EAEC), *mapA* gene of *Campylobacter jejuni, Twhip2* gene of *T. whipplei* and *invA* gene of *Salmonella spp*.

#### Identification of resistance genes

Real-time PCR (qPCR) amplifications were carried out using a C1000 Touch™ Thermal Cycle (Bio-Rad, USA) with the ready-to-use reaction mix ROX qPCR Master according to the manufacturer’s recommendations. The qPCR amplification was used to confirm the presence of (i) ESBL genes: *bla*_CTX-M-A_ and *bla*_CTX-B_ (*bla*_CTX-M_ cluster A and B) and carbapenemase-encoding genes: *bla*_OXA-23_, *bla*_OXA-24_, *bla*_OXA-48_, *bla*_OXA-58_, *bla*_NDM_, *bla*_VIM_, *bla*_KPC_ and (ii) colistin-resistance genes: *mcr*-1, *mcr*-2 (including *mcr*-6), *mcr*-3, *mcr*-4, *mcr-5* and *mcr*-8, by using primers as described and by using specific primers designed in our laboratory (**Table 1**) [16, 21-26].

**Table 1.**
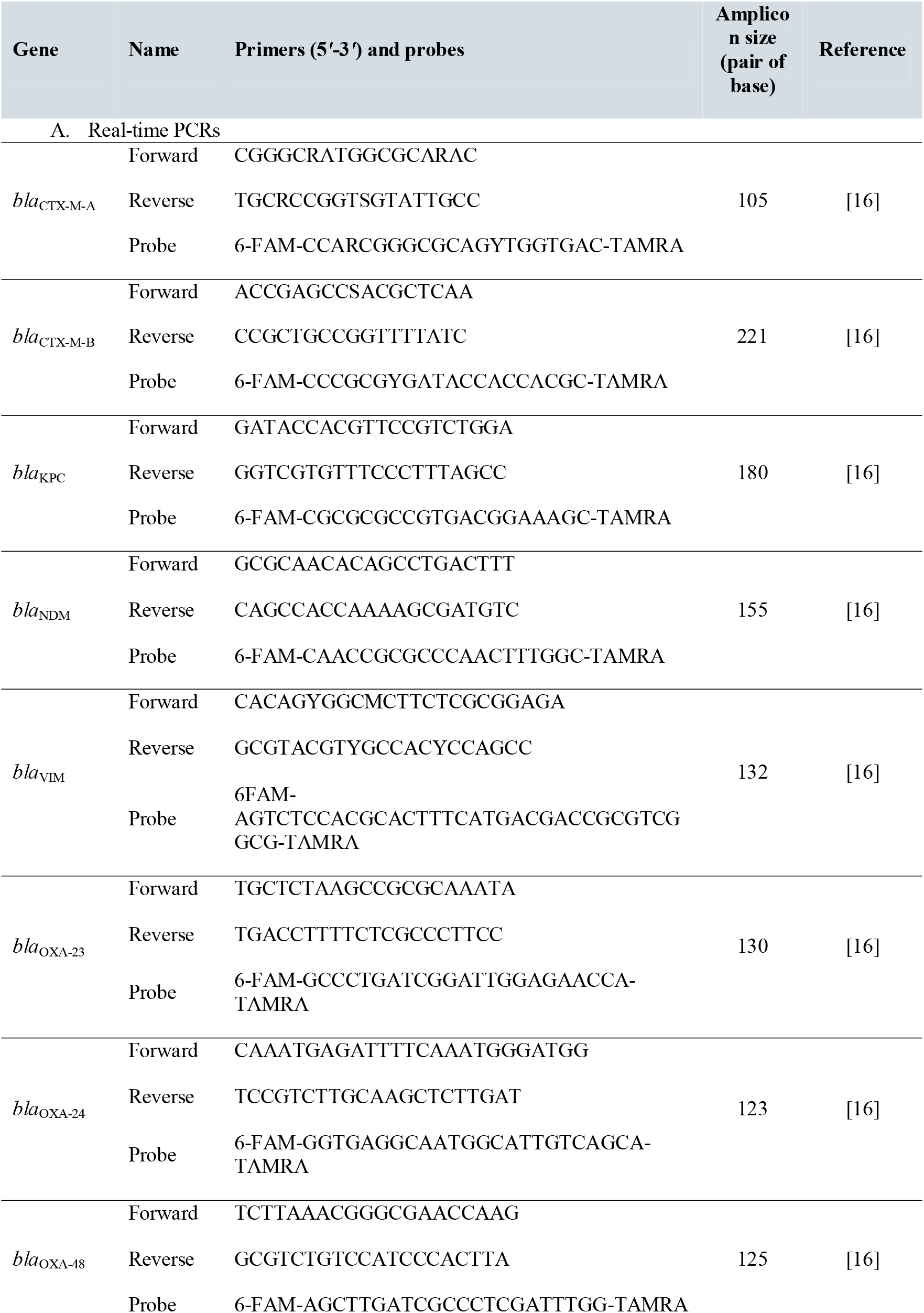
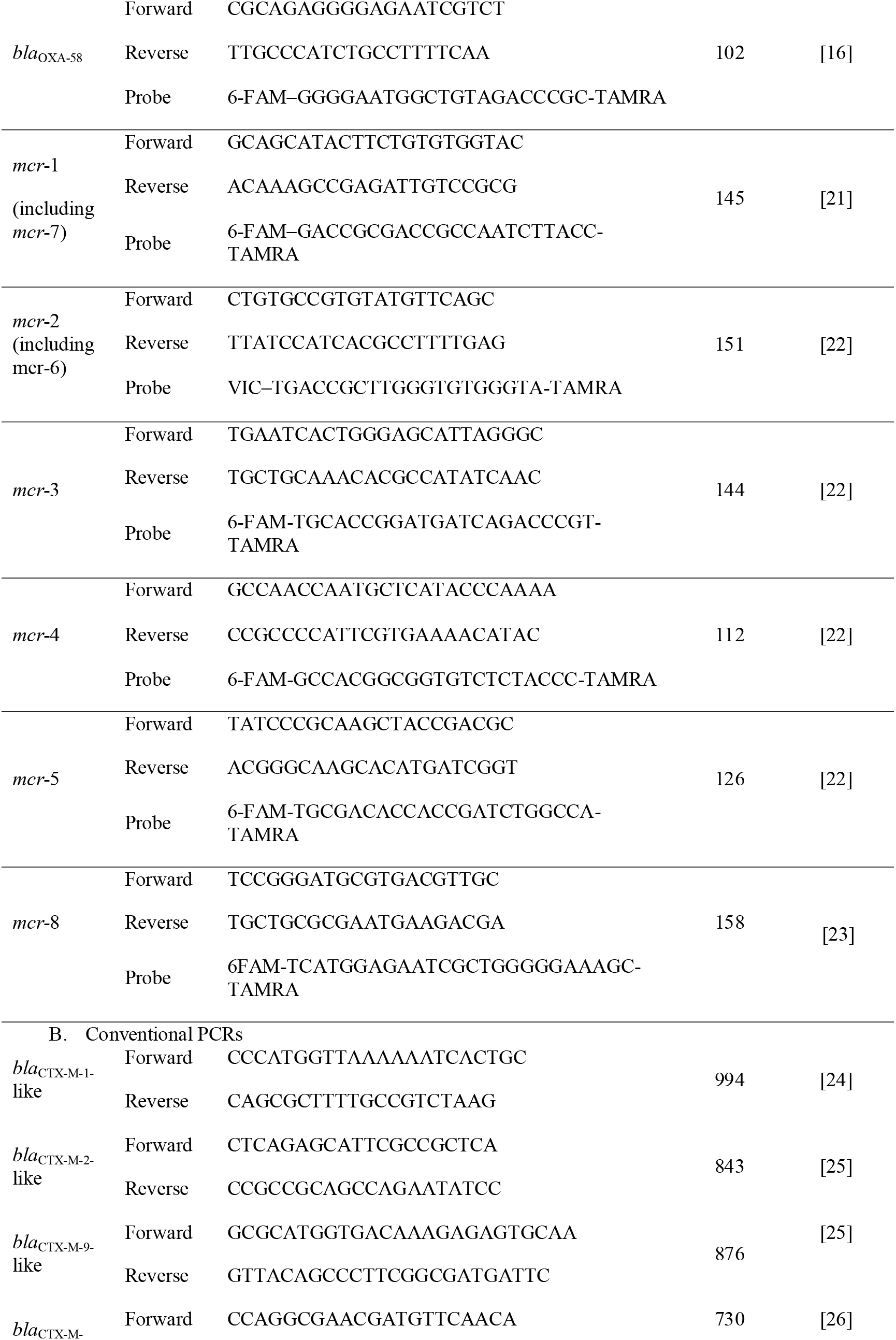
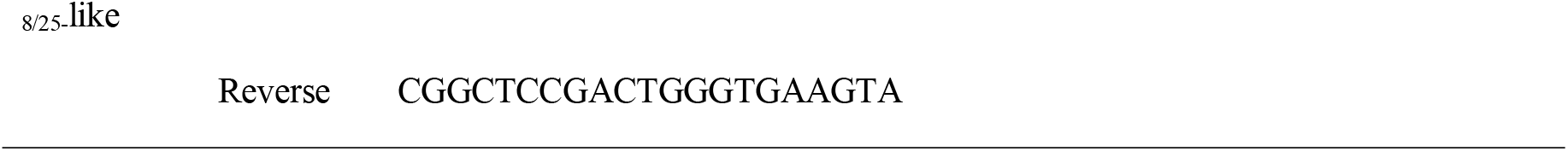
Sequences of primers and probes used for real-time PCRs and conventional PCRs in this study

#### Amplification procedure and experimental validation

Negative control (single PCR mix & sterile H_2_O) and a positive control template (Plasmid DNA extracted from bacterial strains [for enteric pathogens]) or from a colony of cultured *Acinetobacter baumannii, E. coli* or *Klebsiella pneumoniae* [for resistance genes] were included in each qPCR experimental run. Positive results were defined as those with a cycle threshold (CT) value ≤ 35.

### Conventional PCR and sequencing

To better characterize resistance genes, only positive qPCR samples were further tested by conventional PCR. Positive *bla*_CTX-M-A_ samples were tested using two conventional PCR systems for *bla*_CTX-M-1_-like and *bla*_CTX-M-9_-like genes. Positive *bla*_CTX-B_ samples were tested using two conventional PCR systems for *bla*_CTX-M-2_-like and *bla*_CTX-M-8/25_-like genes. The purified positive conventional PCR products were sequenced using specific primers and the BigDye Terminator® version 1.1 cycle sequencing ready reaction mix (Applied Biosystems, Foster City, CA). The sequencing reactions were purified with SephadexG-50 Superfine on MAHVN 45–50 plates (Millipore, Molsheim, France) and then sequenced on the Applied Biosystems 3130 platform (ABI PRISM, PE Applied Biosystems, USA). For each gene, the sequences obtained were edited and assembled using Chromas Pro1.7.7 software (Technelysium Pty Ltd, Australia) and were then aligned with reference genes from the ARG-ANNOT by Mega 7.0 software (https://www.megasoftware.net) [27]. These sequences are available in GenBank at accession numbers **MT215959** and **MT215960** (for *bla*_CTX-M-A_) (**Figure 1**).

**Figure 1.**
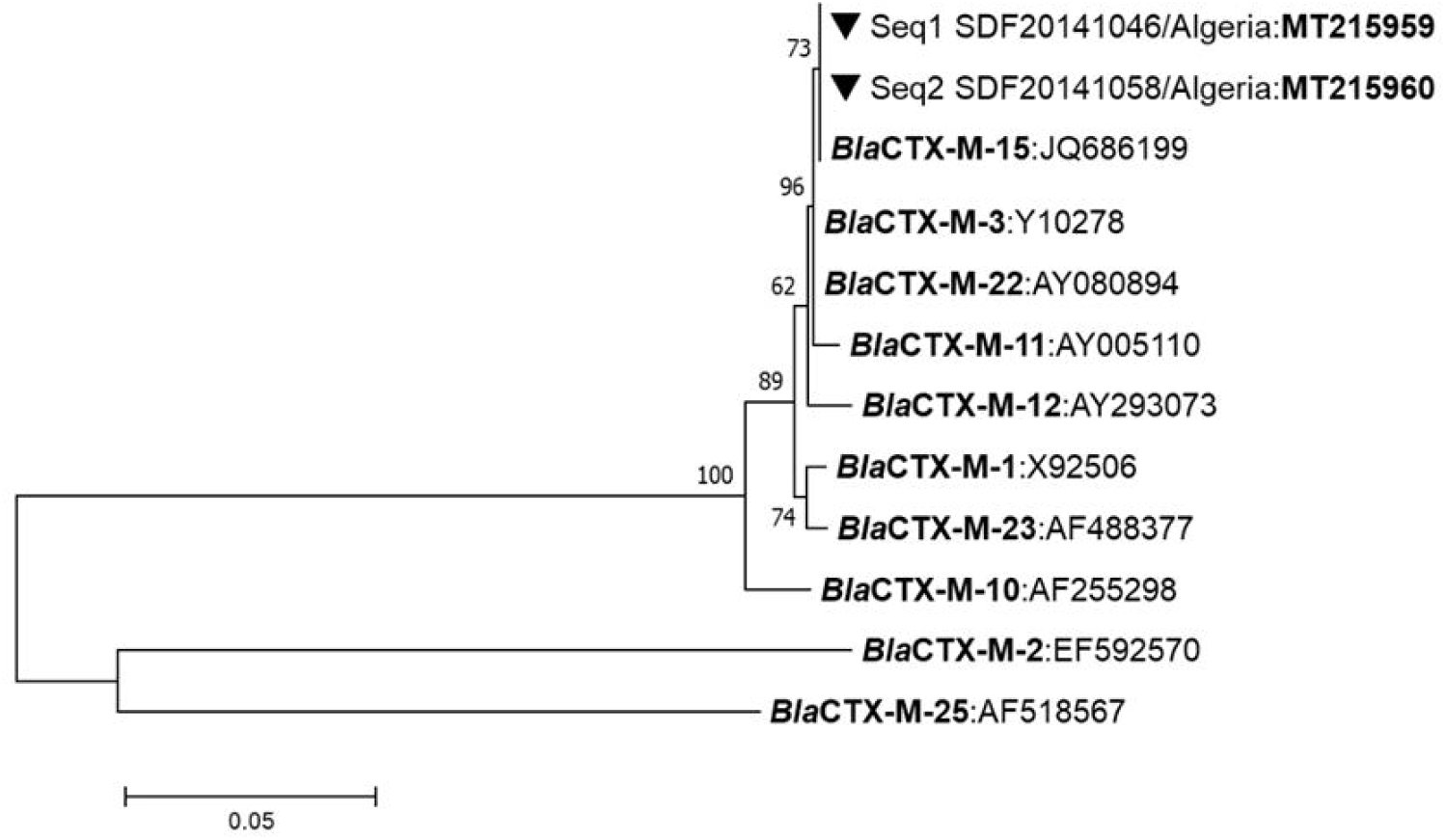
Maximum likelihood phylogenetic tree of the diversity of *bla*_CTX-M_ resistance genes detected in rectal swabs from Marseille homeless people 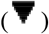. Phylogenetic inferences were conducted in MEGA 7 using the maximum likelihood method based on the Tamura-Nei model.

### Statistical analysis

Statistical procedures were performed using STATA 11.1 software (StataCorp LLC, USA). Statistical differences in baseline characteristics were evaluated by Pearson’s chi-square or Fisher’s exact tests as categorical variables. A two-tailed p-value <0.05 was considered as statistically significant. The odds ratio (univariate analysis) was used to examine associations between the presence of bacterial pathogen DNA and enteric symptoms or migrant status.

## Results

### Population characteristics (Table 2 and Figure 2)

Overall, 114 homeless persons were included in the study and provided rectal samples. The homeless individuals were predominantly middle-aged males, mostly originating from North Africa who settled in France approximately 10 years before the survey was done. The mean duration of homelessness was about 4 years. Overweight status was reported in 48 individuals (44.9%), and obesity in 8 individuals (7.5%).

**Table 2.**
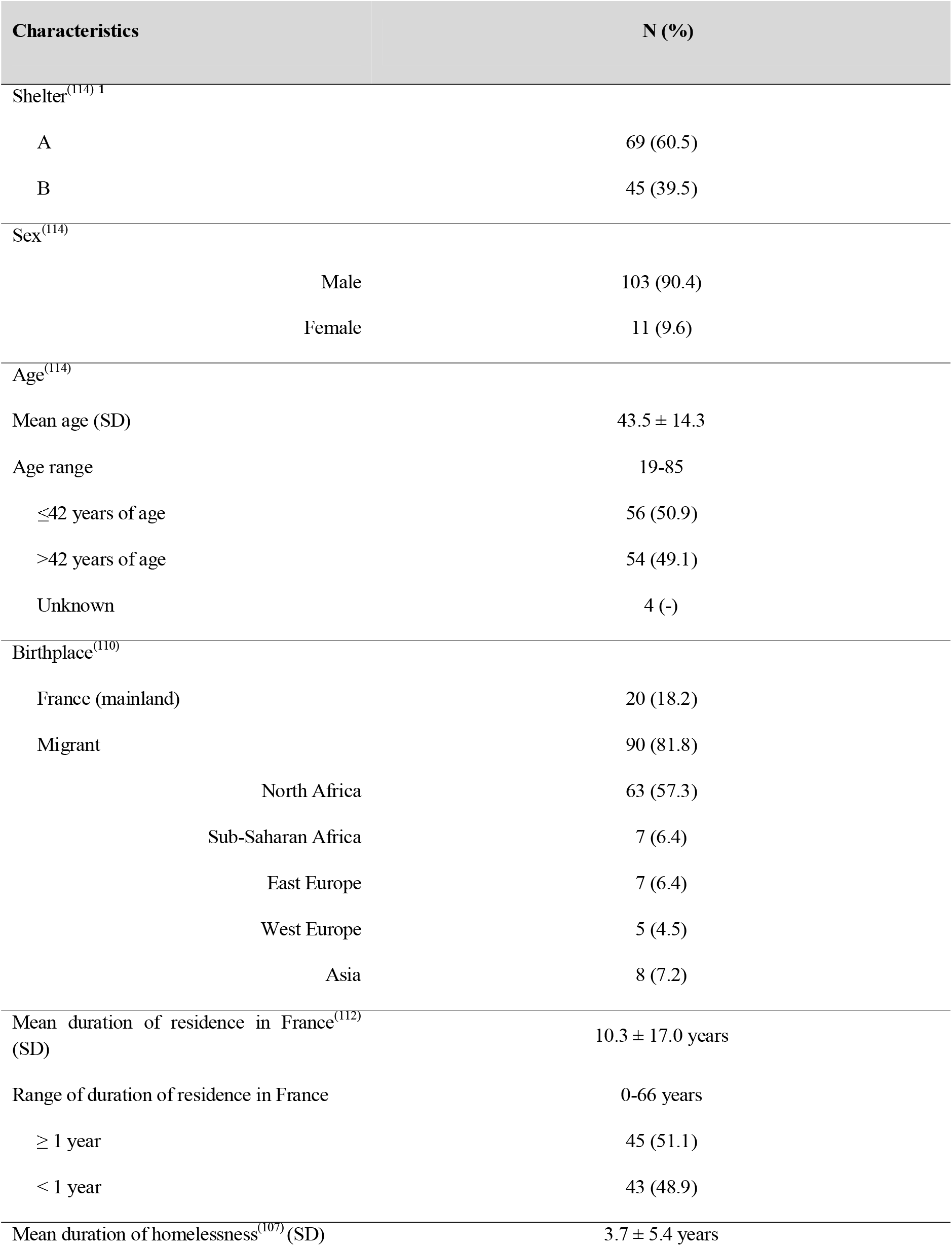
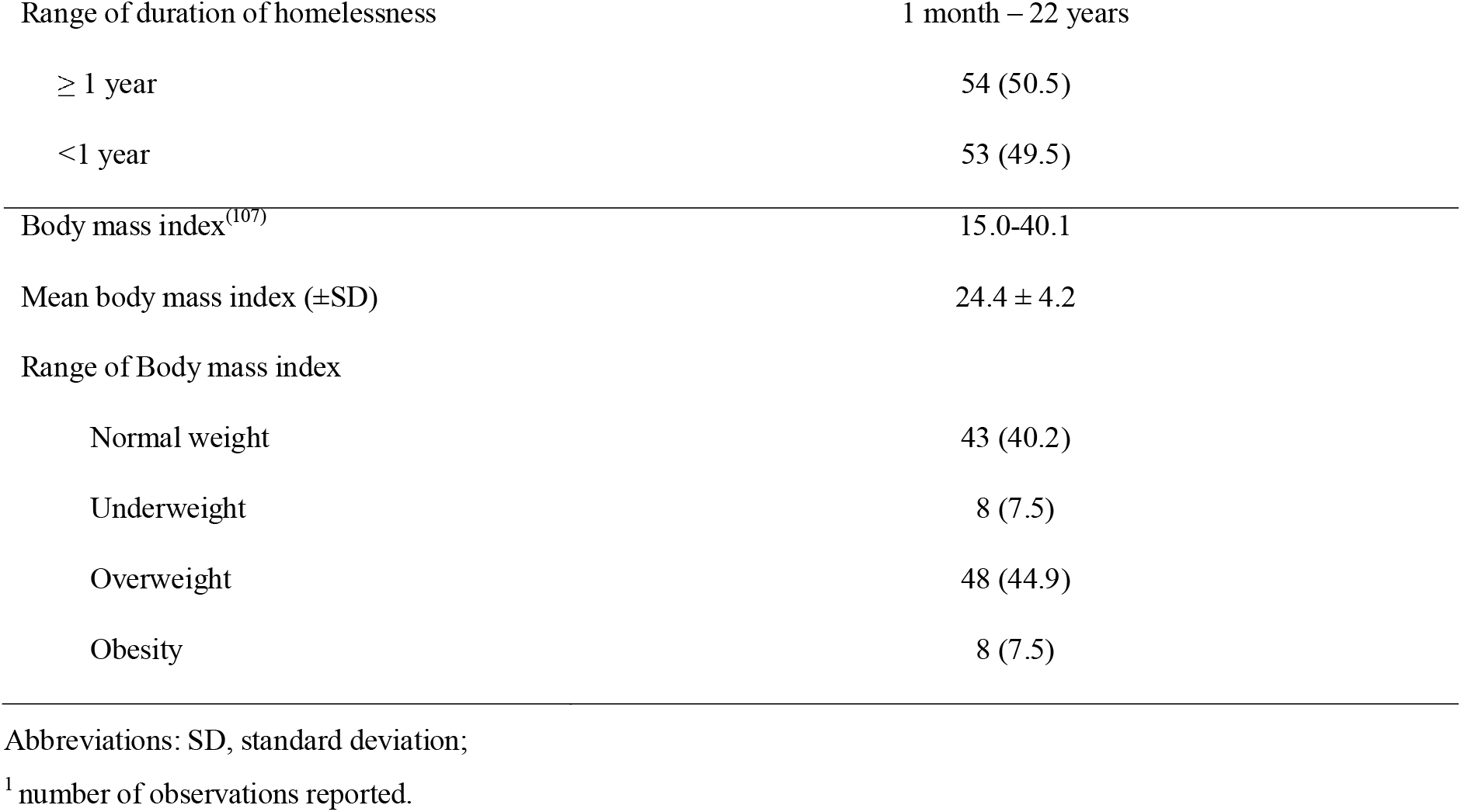
Demographics and body mass index (N=114 individuals)

**Figure 2.**
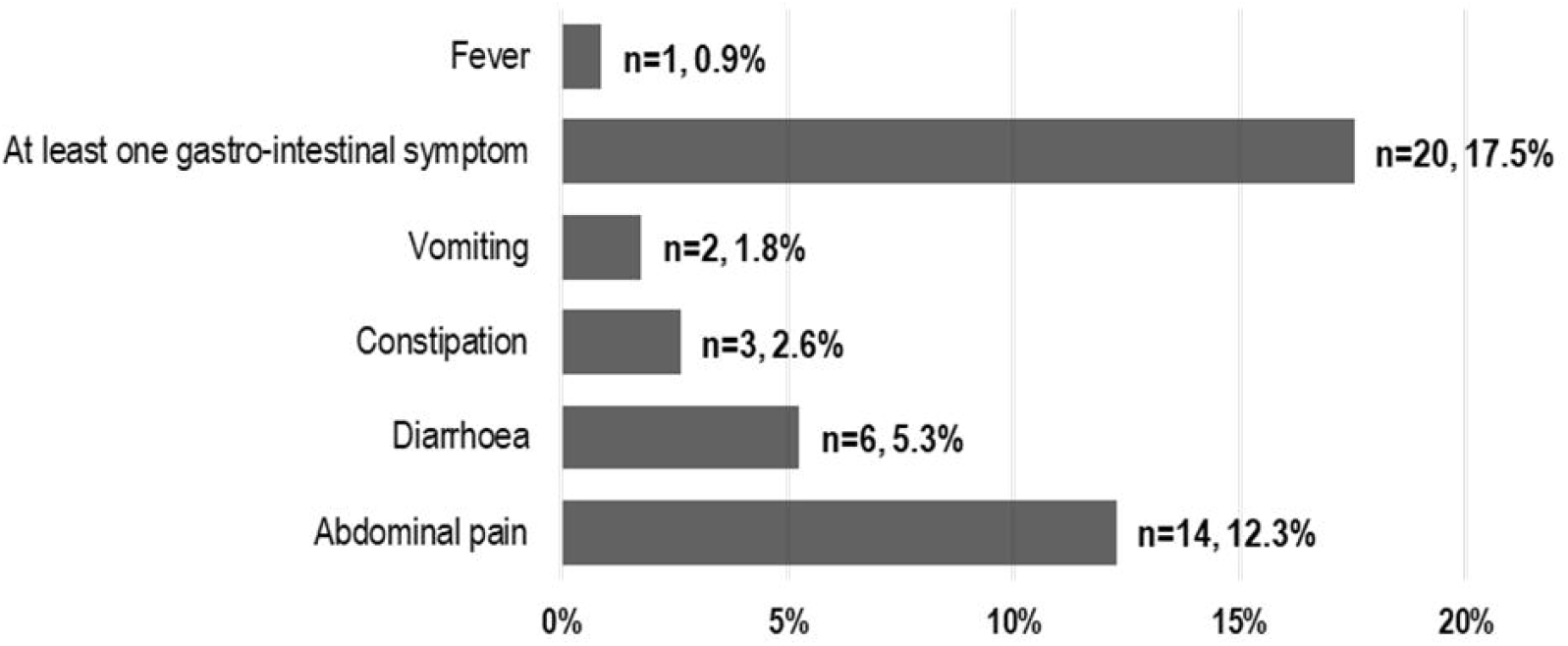
Prevalence of gastrointestinal symptoms (N=114 individuals)

About 17.5% (n=20) declared having at least one gastrointestinal symptom at enrolment, with abdominal pain the most frequent. One participant (0.9%) had fever. None was hospitalized.

### Screening for enteric bacteria (Table 3)

We recorded a 9.6 % prevalence of rectal carriage of bacterial DNA (n=11), with EHEC and EPEC the most frequent. Of positive individuals, seven were born in Algeria, two in France, one in Italy and one in Pakistan. No significant association between carriage of pathogens and gastrointestinal symptoms (odds ratio=1.9 [0.5-7.9], p=0.4) or being a migrant (OR=1.0 [0.2-5.0], p=1.0) was found.

**Table 3.**
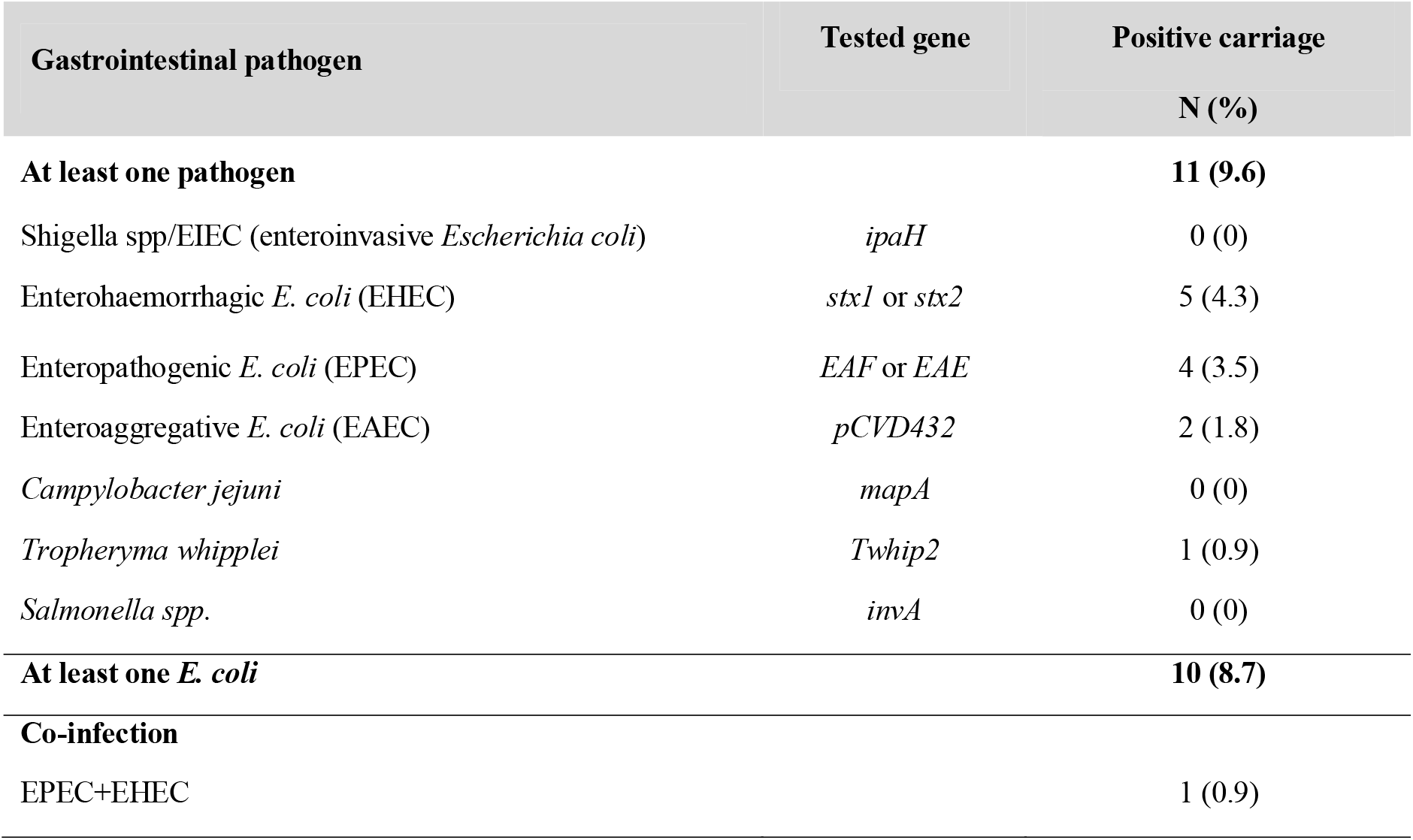
Prevalence (%) of gastrointestinal pathogen DNA detected by qPCR (N=114 individuals)

### Screening for resistance genes (Table 4 and Figure 1)

Only two individuals (1.8%) were positive for *bla*_CTX-M-A_ (qPCR). These two individuals tested negative for bacterial pathogen DNA. Both individuals were further positive for *bla*_CTX-M-1_-like genes (conventional PCR). Both were recruited from shelter A, were from Algeria, having been in France for less than six months and having experienced homelessness for less than five months. These two sequences showed 100% nucleotide identity to *bla*_CTX-M-15_-like type/reference gene at the ARG-ANNOT site.

**Table 4.**
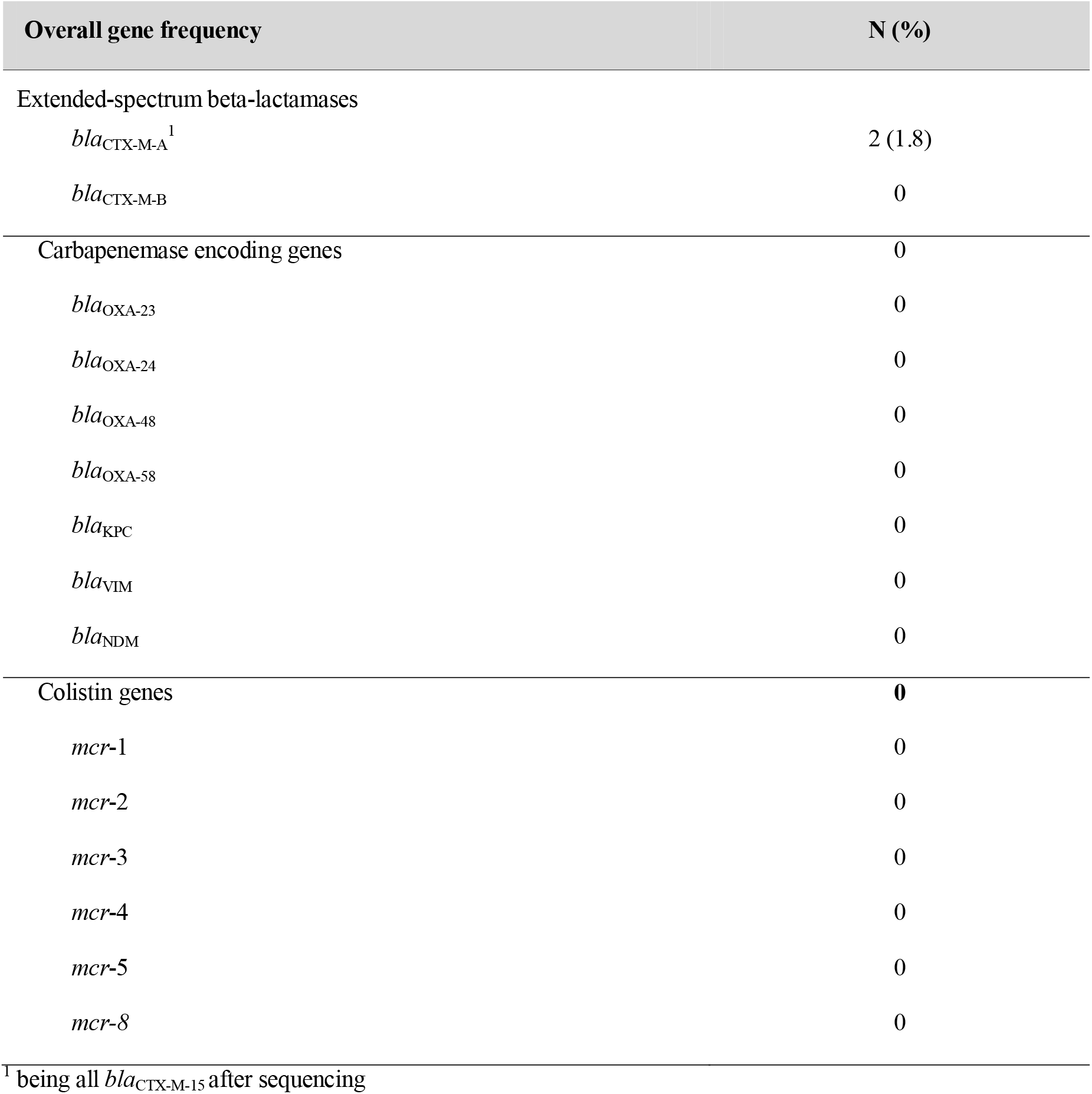
Prevalence of antibiotic resistance genes in rectal samples (N=114 individuals)

| Overall gene frequency | N (%) |
| --- | --- |
| Extended-spectrum beta-lactamases |  |
| *bla*<sub>CTX-M-A</sub><sup>1</sup> | 2 (1.8) |
| *bla*<sub>CTX-M-B</sub> | 0 |
| Carbapenemase encoding genes | 0 |
| *bla*<sub>OXA-23</sub> | 0 |
| *bla*<sub>OXA-24</sub> | 0 |
| *bla*<sub>OXA-48</sub> | 0 |
| *bla*<sub>OXA-58</sub> | 0 |
| *bla*<sub>KPC</sub> | 0 |
| *bla*<sub>VIM</sub> | 0 |
| *bla*<sub>NDM</sub> | 0 |
| Colistin genes | **0** |
| *mcr-1* | 0 |
| *mcr-2* | 0 |
| *mcr-3* | 0 |
| *mcr-4* | 0 |
| *mcr-5* | 0 |
| *mcr-8* | 0 |

<sup>1</sup> being all *bla*<sub>CTX-M-15</sub> after sequencing

None of the samples tested positive for *bla*_CTX-M-B_, carbapenemase-encoding genes and colistin-resistance genes.

## Discussion

This is the first retrospective study aiming to assess the carriage of a panel of bacterial pathogen DNA and resistance genes in rectal samples from sheltered homeless persons in Marseille. We found relatively low rates of *E. coli* pathotype genes (8.7%). The prevalence of *E. coli* DNA carriage was 24.2% among pre-Hajj pilgrims (before departing from Marseille) in the summer between 2016-2018 (unpublished data) [28] and 13.5% among medical students before travelling abroad in the summer 2018-2019 (unpublished data).

One subject only tested positive for *T. whipplei* (0.9%), which is much lower than the 12.9% prevalence observed in 2010-2011 in Marseille sheltered homeless stool samples [14]. A possible explanation for the low rates of pathogen DNA in the present study is that it was carried out in winter, since it was demonstrated that seasonal variations have impacted the enteric microbial community in adults and children [29-30]. Future studies will be conducted at least twice a year (in winter and in summer) to challenge this hypothesis.

Overall, a very low prevalence of resistance gene carriage was observed among homeless individuals, with only two individuals (1.8%) carrying bla_*CTX-M-15*_ genes. In surveys conducted among French pilgrims before departure to the Hajj pilgrimage in the years 2013-2014, the prevalence of bla_*CTX-M*_ was 9.2% when detected in rectal samples by the same molecular method [31]. Our result is in accordance with a low prevalence of resistance gene carriage found in respiratory samples among Marseille homeless persons in 2018. Our work has several limitations. Homeless participants were not randomly selected, so that those harbouring gastrointestinal symptoms might have been more prone to enroll in the survey, given that a medical examination was offered. Viral and parasite pathogens were not tested in this study. Information about recent antibiotic use prior to testing was not documented. The detection of resistance genes directly from rectal samples did not allow identifying the bacteria that housed the antibiotic resistance genes. Despite these limitations, this preliminary study evidenced a relatively low rate of both gastrointestinal pathogen DNA and resistance gene carriage among sheltered homeless persons in Marseille, suggesting that the homeless do not have a high risk of harbouring gastrointestinal antibiotic resistant bacteria.

## Data Availability

GeneBank: accession numbers MT215959 and MT215960 (for blaCTX-M-A)

## Acknowledgments

This study was supported by the Institut Hospitalo-Universitaire (IHU) Méditerranée Infection, the National Research Agency under the program “Investissements d’avenir”, reference ANR-10-IAHU-03, the Région Provence Alpes Côte d’Azur and European funding FEDER PRIMI. We thank our colleagues for their technical assistance.

## Authors’ contribution

TD, JMC and PG contributed to experimental design, data analysis, statistics, interpretation and writing. TD, VT, TL, SB, HTD administered questionnaires, examined patients and collected samples. LH, ML provided technical assistance. DR, JMR contributed to critically reviewing the manuscript. PG coordinated the work.

